# Parkinson’s disease progression and fronto-temporal disconnection relates to identity of minor hallucinations

**DOI:** 10.1101/2025.06.27.25330420

**Authors:** Neza Vehar, Jevita Potheegadoo, Nathalie Heidi Meyer, Léa Florence Duong Phan Thanh, Cyrille Stucker, Marie Elise Maradan-Gachet, Herberto Dhanis, Lucas Burget, Sara Stampacchia, Fosco Bernasconi, Olaf Blanke

**Affiliations:** Laboratory of Cognitive Neuroscience, Neuro-X Institute, School of Life Sciences, Ecole Polytechnique Fédérale de Lausanne (EPFL), Geneva, Switzerland; Department of Neurology, Inselspital, University Hospital Bern, University of Bern, Bern, Switzerland; Department of Clinical Neurosciences, Geneva University Hospital, Geneva, Switzerland; Graduate School for Health Sciences, University of Bern, Bern, Switzerland

**Keywords:** presence hallucination, self-other, phenomenology, resting-state fMRI, social familiarity

## Abstract

Presence hallucinations (PH), non-veridical perceptions of someone appearing around you, are frequently occurring symptoms, in Parkinson’s disease (PD). They belong to the group of minor hallucinations and are predictors of the later appearance of structured visual hallucinations and PD psychosis. Despite their elevated frequency and negative clinical outcomes, differences in PH subtypes have not been examined.

**Objectives:** Investigating PD patients with identified versus unidentified PH, this study determines differences in disease progression, motor and cognitive performance, and brain mechanisms.

**Methods:** Based on detailed interviews of 77 PD patients, 27 reporting PH were grouped into those with identified PH versus unidentified PH (iPH-PD, n=12; uPH-PD, n=15). Disease progression (UPDRS, Hoehn Yahr), cognitive functions (PD-CRS), sensitivity to robot-induced PH and resting-state functional connectivity were analysed.

**Results:** iPH reports mostly consisted of first-degree close others (e.g., partner). iPH-PD patients had a more advanced PD stage, reported more daily-life motor impairments and more simple visual illusions than uPH-PD patients. Both groups showed similar sensitivity to robot-induced PH. Resting-state functional connectivity showed that iPH-PD patients vs. uPH-PD patients had right IFG-pMTG functional disconnection within a previously defined PH-network and the degree of this disconnection was correlated with posterior cognitive impairment.

**Conclusion:** Distinguishing iPH-PD from uPH-PD patients, we show that the former display worse daily motor functioning, more frequent visual illusions and stronger fronto-temporal disconnection, related to cognitive dysfunction. These data suggest that iPH-PD have a more advanced form of PD, possibly representing an intermediate disease stage, between uPH-PD and PD patients with structured visual hallucinations.

## Introduction

Sudden perceptions of intangible presences appearing in one’s vicinity (termed presence hallucinations, PH), are reported in many neurological and psychiatric disorders(1–3), but are particularly prominent in Parkinson’s disease (PD)(4) and Dementia with Lewy bodies (5). Even though 30-45% of PD patients experience PH(3,6–8), differences in phenomenology (ranging from amorphous shadows to fully identified significant others) and its behavioural and brain mechanisms remain poorly understood, despite recent interest in minor hallucinations (including PH, passage hallucinations, visual illusions)(9–12). More research on PH in PD is needed because (1) hallucinations in PD are strong predictors for hospital placement(13), higher cognitive decline(14), decreased quality of life(15), worse disease progression(11) and higher mortality(16); (2) minor hallucinations are associated with more severe psychotic symptoms(17), for example structured visual hallucination(6,18,19) and because (3) PH has recently also been proposed as prodromal diagnostic symptom, presenting as early PD non-motor symptoms, detectable before any motor symptoms(6).

Analysis of hallucination content in clinical research has been carried out previously(20,21), aiding differential diagnostics for example between Alzheimer’s disease and Dementia with Lewy bodies(22) or between bipolar disorder and schizophrenia(23). Linking differences in the phenomenology of hallucinations to stages of disease severity has received most interest in schizophrenia(24,25) while in PD the focus was the distinction between minor versus structured hallucinations, given that patients with the latter hallucinations are more cognitively impaired and at a more advanced stage of the disease(10,12), associated with dementia and depression(26,27). Phenomenologically, approximately 70-88% of PH are perceived as bodies of anthropomorphic human agents(7,8,19) and less frequently as animals or shadows. Moreover, studies suggested that approximately 40-50% of PH in PD are experienced as a person whose identity is familiar to the patient(4,7,8). Such identified PH (iPH) are often of a close relative or partner(6). The remaining PH lack such identity attributes and have sparse phenomenological attributes (e.g., age) or none at all (unidentified PH, uPH). This distinction was first observed in the seminal work by Fénelon(4), who proposed that PH is “a social hallucination”, resulting from neurodegenerative interference with social and person-related posterior brain mechanisms (i.e., autobiographical memory networks). A different line of recent work confirmed that PH are associated with fronto-temporal brain mechanisms(28–31), linking PH to somato-motor mechanisms. This was supported by the experimental induction of PH, using a robotic set-up applying conflicting spatiotemporal signals(32) in healthy individuals(30,33). PD patients were found to be highly sensitive to this procedure(29). Neuroimaging work established a frontotemporal network for PH consisting of six regions: bilateral inferior frontal gyrus (IFG, or ventrolateral PFC), ventral premotor cortex (vPMC) and posterior middle temporal gyrus (pMTG)(29,30,33). However, to date clinical data and brain mechanisms of PD patients with iPH (iPH-PD) versus uPH-PD have not been compared directly.

Here, we examined the clinical relevance of iPH versus uPH, by examining (1) the phenomenology of PH in detail. Critically, we investigated whether 27 PD patients with iPH (iPH-PD, n=12) vs. uPH (iPH-PD, n=15) would differ in (2) motor symptoms(34), cognitive symptoms (fronto-subcortical and posterior sub-scores)(35), overall PD progression stage(36,37), and sensitivity to robotic PH-induction(29,32,33). To probe potentially different brain mechanisms, we (3) investigated iPH-PD vs. uPH-PD differences in resting-state functional connectivity (FC) within a previously defined PH-network(29).

## Methods

### Participants

Seventy-seven PD patients were part of an ongoing clinical study (clinicaltrial.gov pre-registration ID: NCT04579887). From these, we recruited 27 with spontaneous PH (10 women, age mean±SD=69.8±8.6 years; disease duration=9.8±4.6 years). Patients’ PH descriptions from semi-structured interview were used for group classification. If at least one of their PHs could be identified from personal experience (e.g., their partner) or knowledge (cultural, religious context), they were categorised as iPH-PD (n=12, 5 women; age mean±SD=70.7±9.6; disease duration=9.8±5.6). If presences were not recognizable by the patient (e.g. “it just feels like someone there”), or had only partial attributes (e.g., “it feels like an unknown young man”), they were considered uPH-PD (n=15, 5 women; age mean±SD =69.1±8.0; disease duration =9.8±4.0).

Patients underwent neurological examination, signed an informed consent and were included if they reported PH and if this hallucination was present >1x in the past month and/or was recurrent since PD onset. Exclusion criteria involved dementia (Montreal Cognitive Assessment (MoCA) score<18), neurological and psychiatric illness, severe physical disability (including tremors, dyskinesia, akinesia), substance abuse, family history of psychiatric disorders, pharmacological studies participation or presence of factors preventing magnetic resonance imaging (for MRI only).

Due to deep brain stimulation implants only 10 iPH-PD (5 women; age mean±SD=70.6±10.5; disease duration=8.4±4.9) and 12 uPH-PD (3 women; age mean±SD=69.3±8.92; disease duration=9.50±3.73) patients underwent MRI.

### Clinical evaluation

Patients’ levodopa medication and its equivalent daily dose (LEDD)(38), education, symptom onset side and handedness were recorded. They underwent neuropsychological testing with Parkinson’s Disease Cognitive Rating Scale (PD-CRS), consisting of fronto-subcortical and posterior-cortical subscales - sensitive to more severely altered processes progressing to dementia(35). Patients were assessed for Parkinsonism severity with Unified Parkinson’s Disease Rating Scale (MDS-UPDRS: I-IV)(34) and the modified version of Hoehn and Yahr’s (HY) disease stage score(36,37).

Delusions, thought disorders and hallucinations were evaluated using the Scale for Assessment of Positive Symptoms (SAPS)(39). Given that SAPS-Hallucinations subscore does not distinguish between all hallucination types, the presence of visual illusions (VI) and passage hallucinations (PA) respectively was assessed during the interview while visual hallucinations (VH) were assessed with SAPS’ item 7.

### Robot-induced PH procedure

On Day 2, all but two patients (unable to participate due to excessive tremor), performed a somato-motor robotic task shown to induce PH (details in (29,32). Such PH-induction involves a conflict between hand actions performed with the front robot and corresponding feedback on participant’s back, which are received synchronously (no delay between the robots) or asynchronously (robots acting with various delays).

The manipulation started with Task 1 (2-minute of synchronous [0 ms] or asynchronous [500 ms delay] stimulation), each followed by lab-tailored PH Questionnaire (PHQ) with 6 items measuring bodily illusions intensity [0-6 scale]. Subsequent Task 2 (Fig.1D) involved three blocks of 12 trials (one trial consisted of stimulation until reaching 10 back-touches) in randomised delays (0/250/500 ms). Patients were asked “Did you feel as if someone was standing behind you or next to you?” and replied verbally “Yes/ No” after each trial.

**Figure 1.**
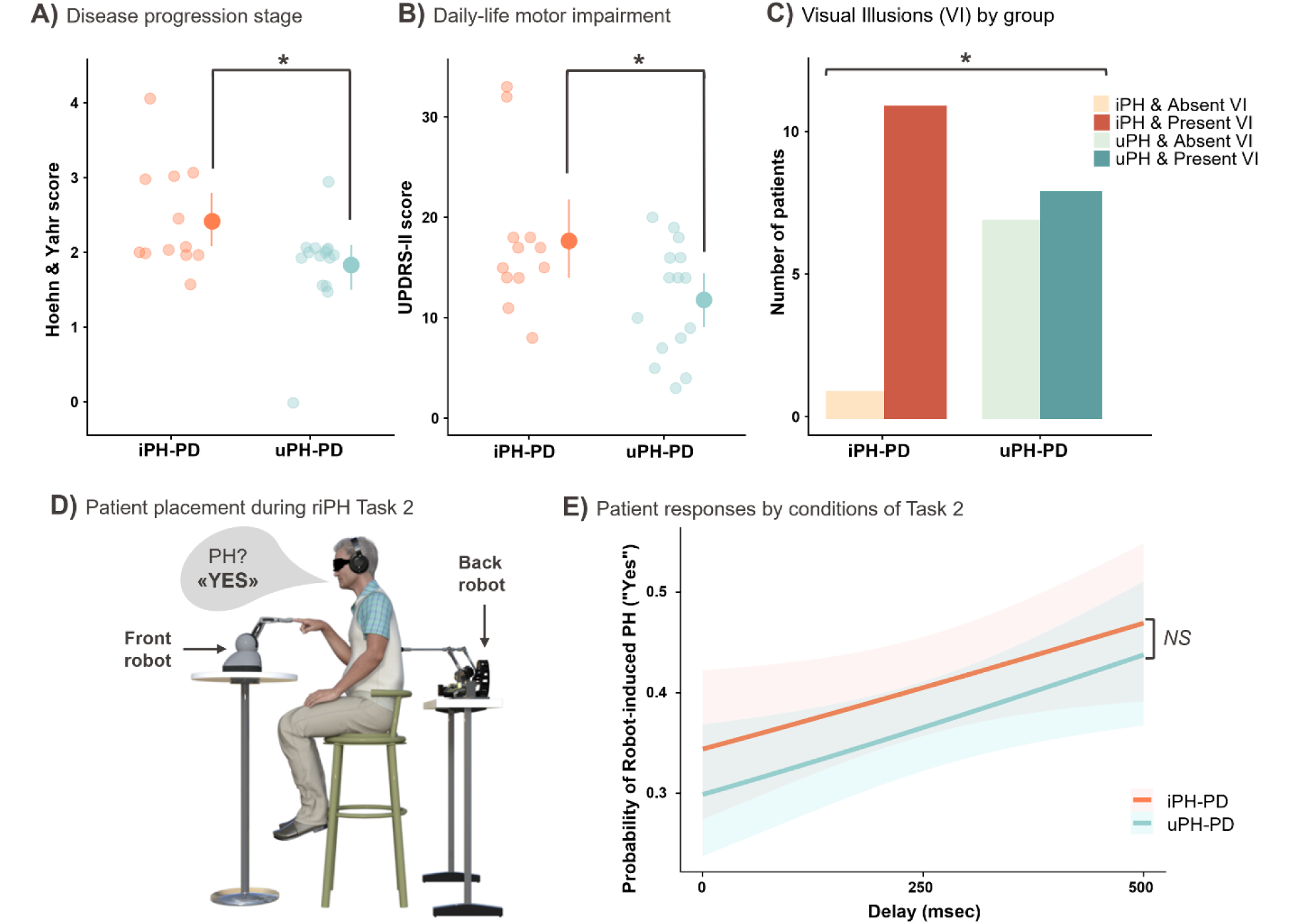
**A,B)** HY and UPDRS-II scores increased for iPH-PD vs uPH-PD. Dots: individuals; dots with bars: group mean, 95% bootstrapped confidence interval. **(C)** Visual illusions, Chi-square p<0.05(*). **D)** riPH set-up. **E)** PH probability linear trend, 95% confidence interval.

### MRI

MRI sequences were acquired with three 3T Siemens MAGNETOM Prisma scanners (Campus Biotech Geneva, Lausanne University Hospital, University Hospital Bern, Switzerland), using 64-channel head and neck coil. We obtained anatomical T1-weighted images, using MPRAGE sequence (slice thickness=1 mm, FOV=256 mm, voxel size=1×1×1 mm, TR=2200 ms, TE=2.98 ms, flip angle=9°, GRAPPA acceleration factor=2). The resting-state scan was acquired with patients having eyes closed. T2*-weighted images were obtained by echo-planar (EPI) BOLD sequences (TR=700 ms, TE=30 ms, flip angle=52°, multiband acceleration factor=8, time points=674, approximate acquisition time=8 minutes)), with an in-plane resolution of 2x2 mm and 2 mm slice thickness (no gap, FOV=256 mm, 72 slices).

MRI analysis was performed using CONN release 22.a(40) and SPM release 12.7771(41). Standard CONN preprocessing (“default_mnifield”) pipeline was used, involving functional realignment with susceptibility distortion correction using fieldmaps, unwarping, slice-timing correction, outlier identification, direct segmentation, normalization, functional smoothing. Acquisitions with framewise displacement above 0.9 mm or global BOLD signal changes above 5 SD were flagged as potential outliers. We normalised slices to the MNI space (voxel size of 2x2x2 mm) and performed spatial smoothing (FWHM=6 mm). Functional data was denoised, by including white matter and cerebrospinal fluid timeseries (16 CompCor noise components) and 2 factors for linear trends within each functional run as confound regressors. Simultaneous bandpass frequency filtering of the BOLD timeseries between 0.008–0.9 Hz was applied. No patients were excluded due to excessive movement (threshold<15% of outlier scans(42).

ROI-to-ROI connectivity matrices (first-level analysis) were estimated characterizing patterns of functional connectivity (FC) with PH network’s 6 regions(29): posterior middle temporal gyri (pMTG), inferior frontal gyri (IFG) and ventral pre-motor cortices (vPMC) bilaterally. FC was computed as the Fisher-transformed bivariate correlation coefficient between each pair of ROIs’ BOLD timeseries.

### Phenomenological interview

Patients described each of their distinct presences in an interview, to perform analysis per PH case. We compared phenomenological traits by categorising all reported presences by shape, age, PH-patient relationship, degree of proximity (First degree: partner, children, parents, siblings; Second degree: parents-in-law, grandparents, grandchildren, aunts/uncles…; Third degree: personally less-known people; Degree undisclosed: if not detailed), and PH gender, where applicable. Note that proximity degree here is based on post-hoc classifications of consanguinity and affinity(43), rather than patients’ subjective perceptions of closeness.

### Statistical Analysis

#### Behavioural data and clinical variables

Analysis was performed using R Studio (2023.12.1+402, R-4.3.2), using sum contrast coding. Linear regression was used to compare neuropsychology scores, clinical and demographic variables between groups. HY score comparison was analysed using cumulative ordinal regression, more appropriate for limited scale ranges(44). Group differences of presence or absence [1/0] of other hallucinations and the number of PH types reported per patient were compared using Fisher test for count data of small samples or Chi-square test, where appropriate. Distribution of counts within many categories were evaluated using Chi-square Goodness-of-fit test, used when >2 categories are possible. In all analyses, the group served as an independent categorical variable, predicting the continuous or ordinal dependent variables in ANOVA Type-III analysis. Where a strong deviation from normality was observed, group differences were evaluated with a Wilcox nonparametric t-test instead (Tab.1).

**Table 1.** Clinical and demographic data of the sample.

|  | Identified<br>presence<br>hallucinations<br>group<br>(iPH-PD),<br>n=12 | Unidentified<br>presence<br>hallucinations<br>group<br>(uPH-PD),<br>n=15 | Total sample,<br>n=27 | *P*<br>(statistical<br>significance<br>of test at<br><0.05) |
| --- | --- | --- | --- | --- |
| **Age (in years)** | 70.7 ± 9.58 | 69.1 ± 7.95 | 69.8 ± 8.57 | 0.639 |
| **Gender (% of women)** | 5 women<br>(41.67%) | 5 women<br>(33.33%) | 10 women<br>(37%) | 0.706 |
| **Education (in years)** | 16.8 ± 5.50 | 16.9 ± 3.67 | 16.9 ± 4.48 | 0.918 |
| **PD duration (in years)** | 9.83 ± 5.56 | 9.80 ± 3.97 | 9.81 ± 4.64 | 0.986 |
| **Hoehn and Yahr stage** | 2.42 ± 0.702 | 1.83 ± 0.617 | 2.09 ± 0.708 | 0.036* |
| **LEDD dosage (in mg)** | 967 ± 654 | 906 ± 388 | 933 ± 513 | 0.766 |
| **Dopamine agonist equivalent dose (in mg)** | 142 ± 196 | 104 ± 116 | 121 ± 154 | 0.860<sup>†</sup> |
| **First site of symptom onset (lateralisation)** | On the right: 6<br>(50.0%)<br>On the left: 5<br>(41.7%)<br>On both sides: 0<br>(0%)<br>Unknown: 1<br>(8.3%) | On the right: 7<br>(46.7%)<br>On the left: 7<br>(46.7%)<br>On both sides: 1<br>(6.7%)<br>Unknown: 0<br>(0%) | On the right: 13<br>(48.1%)<br>On the left: 12<br>(44.4%)<br>On both sides: 1<br>(3.7%)<br>Unknown: 1<br>(3.7%) | 0.909 |
| **Handedness (%)** | Righthanded: 10<br>(83.33%),<br>Lefthanded: 1<br>(8.33%),<br>Ambidextrous:<br>1 (8.33%) | Righthanded: 15<br>(100%)<br>Lefthanded: 0<br>(0%),<br>Ambidextrous:<br>0 (0%) | Righthanded: 25<br>(92.6%)<br>Lefthanded: 1<br>(3.7%)<br>Ambidextrous:<br>1 (3.7%) | 0.188 |
| **MoCA score** | 26.3 ± 2.86 | 27.1 ± 2.13 | 26.7 ± 2.47 | 0.443<sup>†</sup> |
| **MDS-UPDRS I** | 17.8 ± 6.97 | 16.4 ± 6.10 | 17.0 ± 6.41 | 0.574 |
| **MDS-UPDRS II** | 17.7 ± 7.51 | 11.8 ± 5.61 | 14.4 ± 7.05 | 0.028* |
| **MDS-UPDRS III** | 17.1 ± 7.76 | 22.2 ± 11.70 | 19.9 ± 10.3 | 0.204 |
| **MDS-UPDRS IV** | 5.75 ± 5.58 | 4.20 ± 3.36 | 4.89 ± 4.46 | 0.521<sup>†</sup> |
| **Total PD-CRS score** | 91.9 ± 14.4 | 95.9 ± 16.5 | 94.1 ± 15.4 | 0.513 |
| Posterior cortical<br>subscore | 28.4 ± 1.93 | 29.1 ± 1.39 | 28.8 ± 1.65 | 0.356<sup>†</sup> |
| Fronto-subcortical<br>subscore | 63.5 ± 12.9 | 66.9 ± 16.5 | 65.4 ± 14.8 | 0.567 |
| **Total SAPS score** | 7.25 ± 2.45 | 6.80 ± 5.27 | 7.00 ± 4.19 | 0.788 |
| Hallucinations<br>subscore | 6.50 ± 2.61 | 5.47 ± 4.19 | 5.93 ± 3.55 | 0.463 |
| Delusions<br>subscore | 0.33 ± 0.89 | 1.07 ± 2.28 | 0.74 ± 1.81 | 0.503<sup>†</sup> |
| Bizarre Behaviour<br>subscore | 0 ± 0 | 0 ± 0 | 0 ± 0 | NA |
| Positive Formal<br>Thought Disorder<br>subscore | 0.42 ± 0.80 | 0.27 ± 1.03 | 0.33 ± 0.92 | 0.252<sup>†</sup> |

Note. The values provided are means ± SD, or the category percentage inside the group. Statistical significance of tests was considered at threshold of $P < 0.05^*$ . NA: all the values in the subscale were 0, no statistical tests could be performed. <sup>†</sup> Due to strong deviation from normality, a Wilcoxon t-test for nonparametric data was performed. Categorical variables (gender, handedness, and lateralisation) were compared using Fischer test for small sample size.

For robotic PH-induction, the PHQ ratings were compared between groups employing ordinal regression for each item per condition [0/500ms delay, categorical variable] (cumulative link mixed-effects model with random intercept for patients: PH rating∼Group*Delay+(1|patient_ID)). For the riPH task, logistic mixed-effects model was employed, predicting binomial responses by interaction of Group*Delay [0/250/500 ms as continuous variable, z-scaled to avoid convergence issues], with a random intercept for patients: PH[yes/no]∼Group*Delay+(1|patient_ID).

#### Functional connectivity analysis and cognitive performance

Fisher-transformed bivariate correlation coefficients [z-score] between 6 ROI pairs (15 edges/connections) were yielded to R across the entire sample. To establish which edge was different across the two groups, linear mixed-effects regression was used with connectivity as the dependent variable, Edge-name and Group as fixed effects, and random intercept for patients (FC∼Edge*Group+(1|patient_ID)). The results of post-hoc analyses were corrected for multiple comparisons, using p-FDR<0.05 significance cutoff in R. The groups’ variability of FC strength was compared using Levene’s nonparametric test for homogeneity of variance, and their respective indices of dispersion D were calculated. FC of significantly differing edge was correlated with overall cognitive performance from both groups (PD-CRS’s fronto-subcortical and posterior-cortical subscores), following Bernasconi et al.(29), applying Spearman’s rank correlation to account for non-normality, differences in variance and to reduce the influence of extreme values.

## Results

### Clinical and demographic variables

Groups did not significantly differ in age, education, PD duration, dopamine agonist or levodopa-equivalent daily dose, gender, handedness, or symptom onset lateralisation (Tab.1). Accordingly, these variables were not included as covariates in further analyses.

### Motor performance, cognition and disease progression

The iPH-PD versus uPH-PD group were characterized by a significantly worse impairment of daily-life motor actions (UPDRS-II: β=2.93, 95% CI[0.34,5.53], p=0.028; Fig.1B), with the total model explaining a statistically significant moderate proportion of variance (R^2^=0.18) of results. There was no significant group differences in patients’ overall daily life quality (UPDRS-I: β=0.72, 95% CI[-1.87,3.31], p=0.574), motor impairments (UPDRS-III: β=2.56, 95% CI[-1.48,6.60], p=0.204), or motor complications, such as dyskinesia (UPDRS-IV: W=103.5, 95% CI[-3.00,6.00], p=0.521).

The groups did not differ in their cognitive performance on MoCA (W=74, 95% CI[-2,1.00], p=0.443) or overall PD-CRS (β=-2.01, 95% CI[-8.24,4.22], p=0.513), neither in the posterior-cortical (W=72, 95% CI[-2.00,0.00], p=0.356) nor the fronto-subcortical subscore (β=-1.68, 95% CI[-7.66,4.29], p=0.567).

Concerning disease severity, the odds of having worse disease severity (HY score) were 2.61 times higher for the iPH-PD than the uPH-PD group, a significant group difference (z=2.10, 95% CI[1.14,7.35], p=0.036; Fig.1A). The two groups did not significantly differ in delusion severity (W=79.5, 95% CI[-0.00,0.00], p=0.503) or other positive symptoms measured by the SAPS (Tab.1). There were also no significant differences in passage hallucinations (iPH-PD: 91.7%; uPH-PD: 66.7%; p=0.182) or structured visual hallucinations (iPH-PD: 8.3%; uPH-PD: 40%; p=0.091). However, iPH-PD vs. uPH-PD reported significantly more visual illusions (iPH-PD: 91.7%; uPH-PD: 53.3%; p=0.043, Fig.1C).

### Robot-induced PH

In Task 1, robot-induced PH questionnaires showed an expected main effect of Delay with higher ratings for PH in the asynchronous vs. synchronous condition (OR=2.68, 95% CI[1.06,6.79], p=0.038), replicating past results(29,30). There was no main effect of group (OR=0.47, 95% CI[0.10,2.23], p=0.344) or its interaction with condition (OR=1.25, 95% CI[0.56,2.81], p=0.589) (for details, see Supplemental information).

In Task 2, replicating past results(29) the probability of reporting PH significantly increased with the increase of applied somato-motor conflict (Delay main effect: β=0.36, 95% CI[0.18,0.53], p<.001, explaining substantial R^2^=62% of variance). The groups’ overall bias did not differ (main effect: β=0.002, 95% CI[-0.84,1.04], p=0.835) and no significant interaction was found (Group*Delay: β=0., 95% CI[-0.18,0.18], p=0.984) (Fig. 1E). These data show that both groups were sensitive to the robotic somato-motor procedure and that iPH-PD and uPH-PD showed similar sensitivity.

### PH network’s resting-state functional connectivity

Resting-state fMRI analysis revealed that FC is significantly predicted by a near-significant Edge*Group interaction (F(14,280)=1.71, p=0.052) and Edge (main effect F(14,280)=10.21, p<0.001). There was no group main effect (F(14,280)=1.33, p=0.262). We then performed post hoc analysis, exploring group FC differences by edge (adjusted for multiple comparisons), revealing that FC between right IFG and right pMTG was significantly reduced in iPH-PD vs. uPH-PD (p-FDR=0.020) (Fig.2). No other contrast survived the correction (see Supplementary Information). Right IFG-pMTG FC variability was also significantly different between groups (F=8.46, p=0.009), with uPH-PD group being significantly more heterogenous in their connectivity strength (Index of dispersion D=0.095) than iPH-PD (D=0.055).

**Figure 2.**
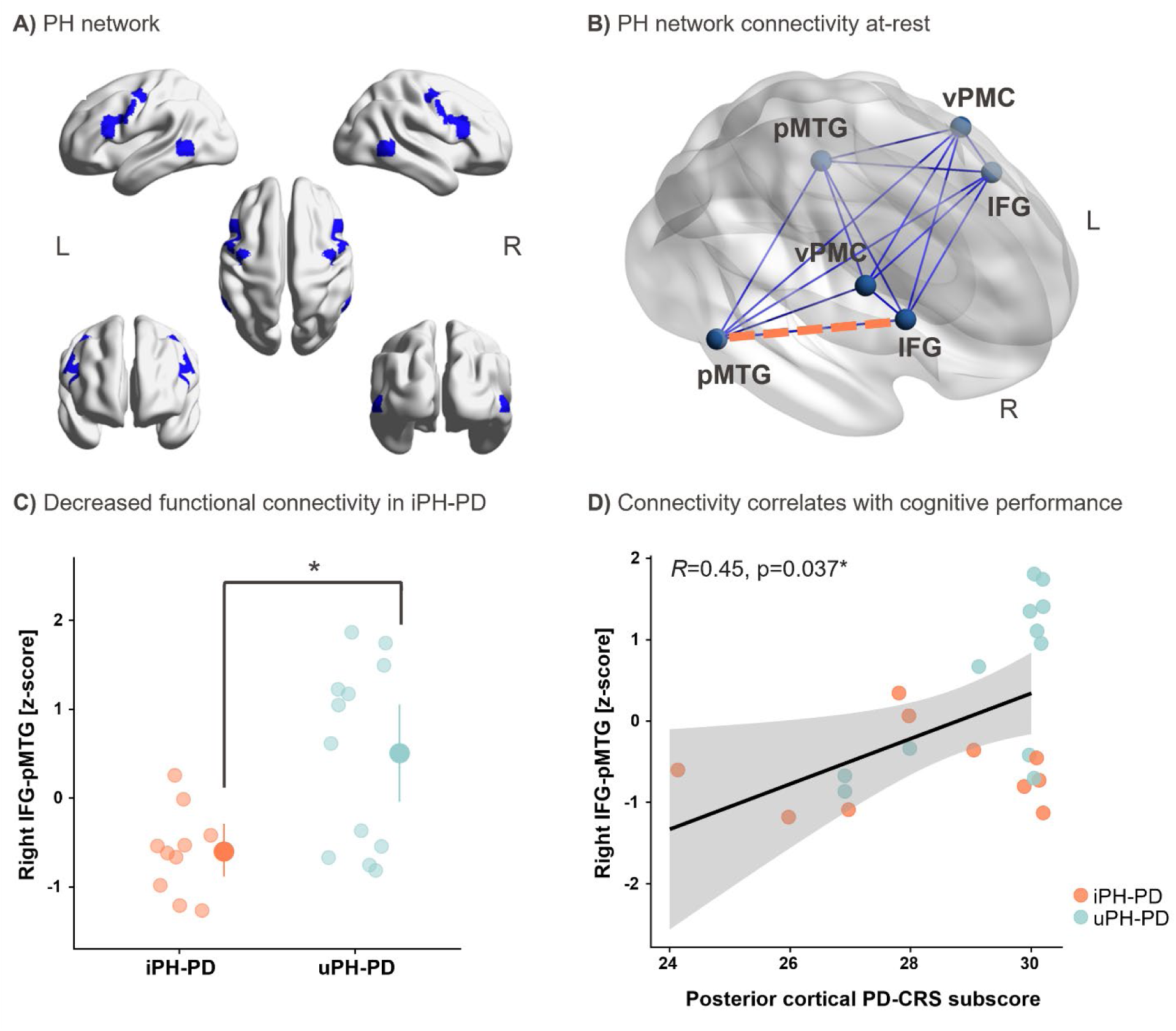
**A)** PH network in MNI space. **B)** Functional connectivity (blue): significantly lower edge for iPH-PD vs uPH-PD (orange). **C)** Individual score and summary dots (mean FC, 95% bootstrapped confidence interval). **D)** Spearman’s correlation: trend line, 95% confidence interval.

Following Bernasconi et al.(14,29), we investigated whether fronto-temporal connectivity is associated with cognitive performance by correlating the altered right IFG-pMTG connectivity with PD-CRS subscores across the two groups (Fig.2D). IFG-pMTG connectivity was significantly positively correlated with posterior cognitive functions (rho=0.45, p=0.037), meaning lower IFG-pMTG connectivity was correlated with a stronger posterior cognitive deficit. No such correlation was found for fronto-subcortical subscore (rho=0.18, p=0.41).

### Phenomenology of Presence Hallucinations

Analysis of interview data showed that individual patients may report a single presence or several distinct cases of PH (range 1-4, mean±SD=1.74±1.06). Patients reported 46 PHs in total, however, the proportion of reported iPH and uPH cases was not significantly different (iPH: 39.9%; uPH: 60.9%; χ²(1)=2.17, p=0.14) in the overall sample. The large majority (82.6%) of reported presences were anthropomorphic (χ²(2)=50.26, p<0.001). Moreover, 73.3% of human iPH cases were recognized as first-degree relatives (χ²(3)=19.4, p<0.001) or closest others (i.e., partner (22.2%), sibling (16.7%)…) (Fig.3, Tab.2).

**Figure 3.**
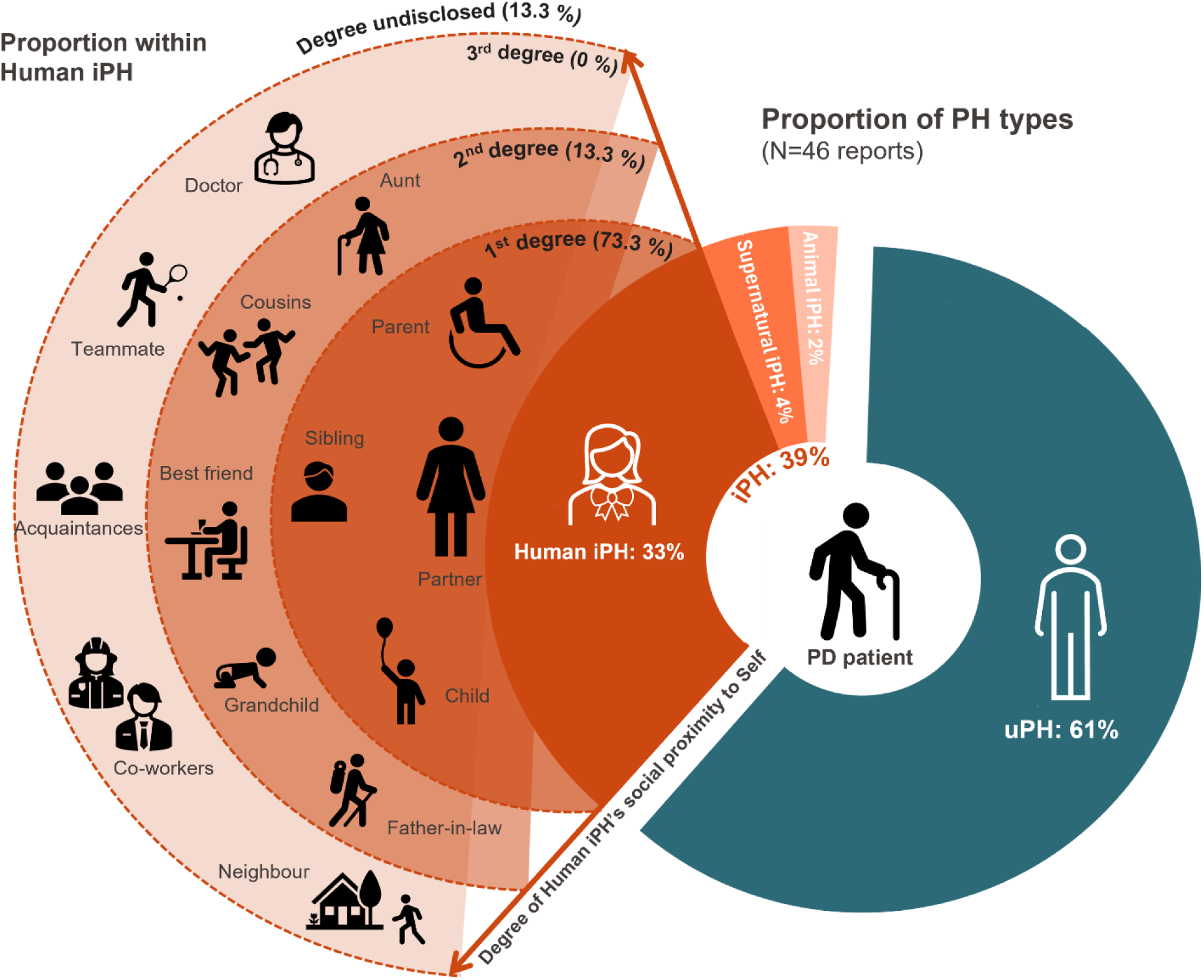
Distribution of PH phenomenology categories in the full sample (27 PD patients): iPH in shades of red, uPH in shades of blue. Concentric circles of the Human iPH represent relationship closeness of the hallucinated person to the patient.

**Table 2.**
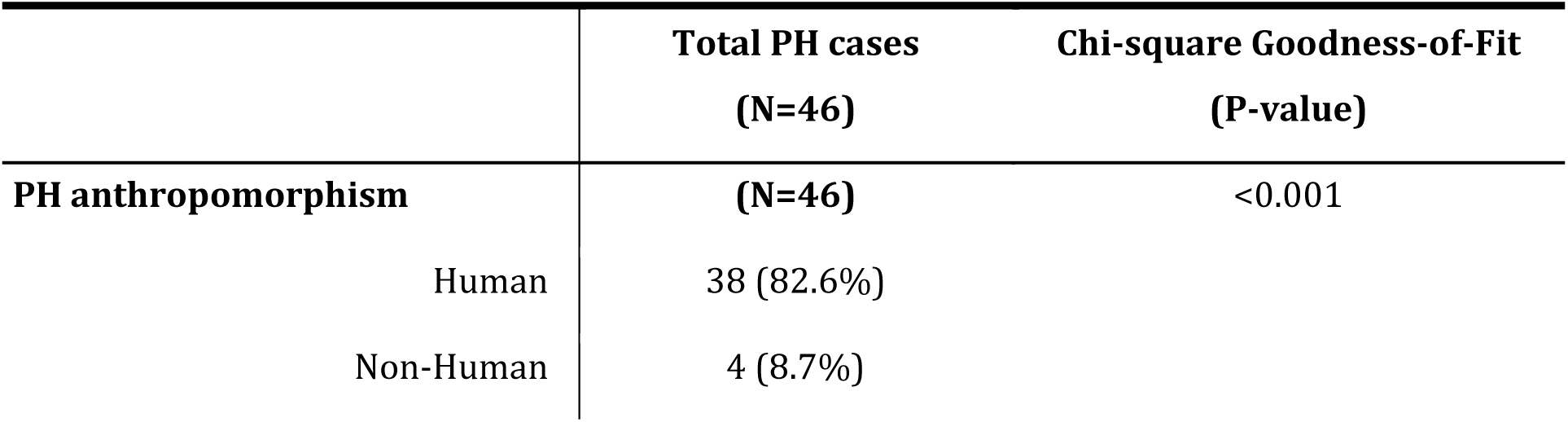
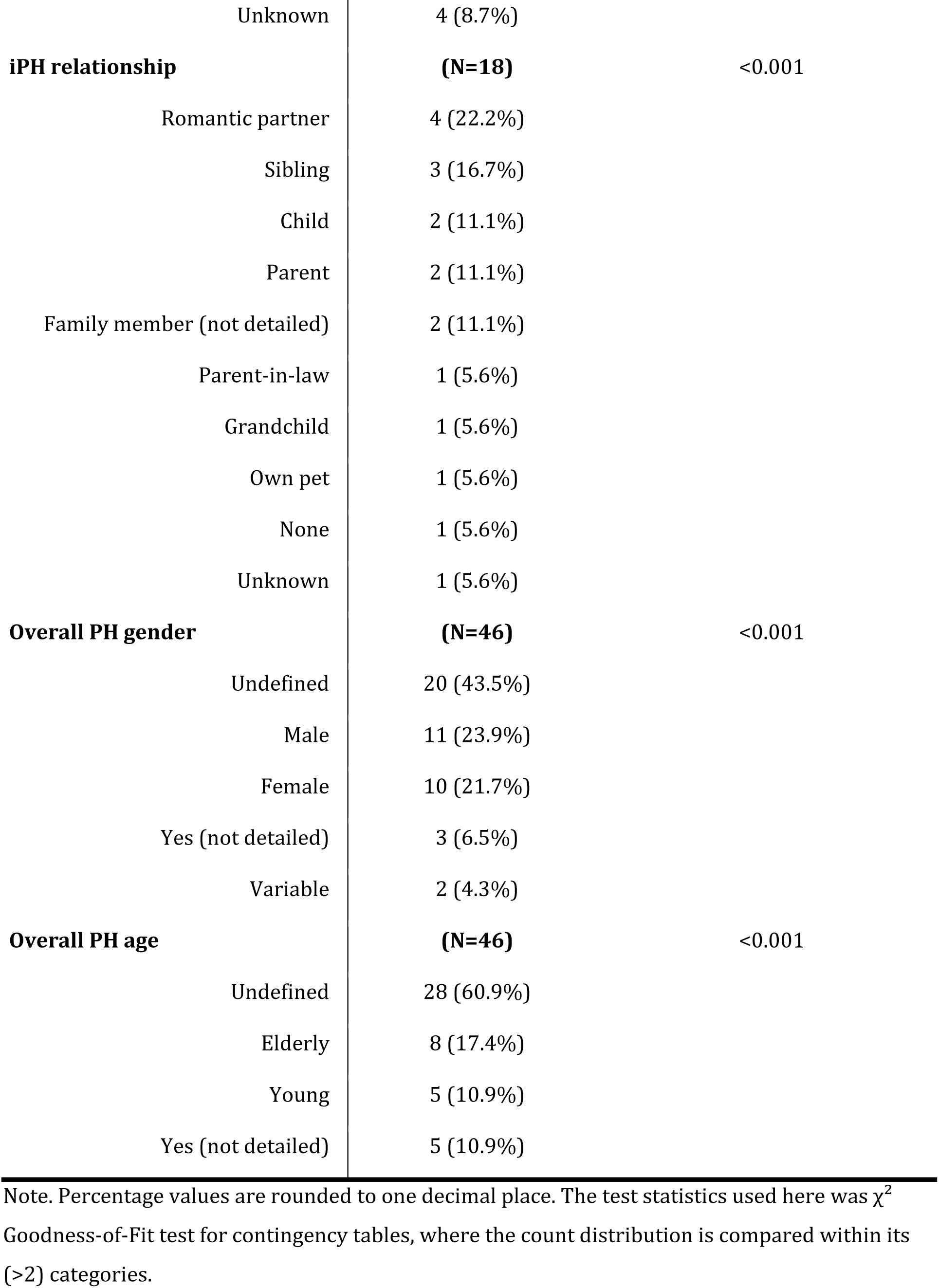
Described PH phenomenology with data distribution significance of within-category representation.

## Discussion

In PD patients reporting PH, we found that (1) the majority of cases consisted of a human presence (83%), of which 73% were recognized as first-degree relatives (for iPH cases). Investigating whether the distinction between iPH-PD vs. uPH-PD is associated with clinical and neural differences, we found that iPH-PD (2) report more severe daily motor impairments (UPDRS-II), are (3) more likely to have visual illusions, and (4) have more advanced PD (HY). These differences were extended by MRI analysis revealing (5) altered fronto-temporal connectivity in iPH-PD, that was further linked to posterior brain functions. We argue that these clinical and neural differences are compatible with more advanced neurodegeneration in iPH-PD vs. uPH-PD, likely reflecting stronger involvement of posterior brain networks, including those processing information about close familiars.

### Hallucinated identity

The present finding of 83% of patients reporting human-like PH is consistent with previous literature indicating that felt presences in PD are predominantly described as anthropomorphic agents (Kataoka & Ueno(19):70%; Wood et al.(8):71%; Reckner et al.(7):88%). Moreover, combining all distinct PH cases that PD patients reported in our sample, 39% were identifiable from one’s personal experience or knowledge (iPH), compared to 61% of unidentifiable (uPH). This result mirrors previous findings estimating 34-42% of PH as “familiar presences” in PD ((4):42%; (8):34%; (7):40%) and healthy population(45). Analysing iPH by social proximity to the patient we found that a significant proportion (73%) is of a first-degree person, most commonly romantic partners and siblings, followed by second-degree (13.3.%) persons (e.g. grandchildren). Interestingly, no patient reported PH of a third-degree person also known to the patient (e.g. co-workers), suggesting that familiarity alone is insufficient for iPH.

### iPH associated with more advanced PD

Previous work in PD highlighted clinical relevance of hallucination phenomenology analysis. For example, differences in disease severity were linked to hallucination complexity (i.e., structured indicating more advanced disease state than minor hallucinations)(9,11,18), sensory modality(3) and insight(46). Here, we investigated whether iPH-PD versus uPH-PD show differences in PD-related impairments. To our knowledge, the only other study investigating hallucination identity content in relation to disease progression and/or cognitive changes in PD was done for structured VH(47), finding worse short-term memory in patients with structured visual hallucinations of a familiar person (akin to identified VH) versus unfamiliar people. The present study extends such data, showing that the iPH/uPH distinction is also clinically useful. Specifically, we found that self-reported impairments in daily motor activities (UPDRS-II) indicated higher motor dysfunction in iPH-PD versus uPH-PD patients. This score has previously been shown to be higher for PD patients with minor versus no hallucinations(48), those with structured VH versus those without hallucinations(49) and for PD patients, who developed psychosis over a 12-year period, compared to those who did not(50). Moreover, iPH-PD versus uPH-PD also had stronger progressive disease-related physical impairments (HY), as quantified by clinicians. Both findings (HY, UPDRS-II) suggest that the present iPH-PD patients suffer from a more advanced PD form, as found for PD patients with complex visual versus minor hallucinations(51). The iPH-PD group also reported significantly more visual illusions than uPH-PD, previously suggested to antedate the development of structured VH.

Based on this, we argue that iPH are instances of fully-fledged identification of the hallucinated person and may accordingly be characterized as a form of structured, albeit non-visual, hallucination. Following this proposal, iPH would represent an intermediate stage between uPH (unstructured, non-visual) on one side and structured visual hallucinations on the other. Structured versus simple PH (iPH vs. uPH) may rely on comparable processes, as were argued in the visual domain for structured versus simple VH(11,17,52,53). Such progression for VH is generally accompanied by worsening of motor and cognitive symptoms(6,11,18,27,54), particularly in attentional and visuospatial dysfunction. The present data suggest a similar progression within PH, with a more advanced disease progression for iPH-PD, even though the present groups are comparable in age, dopaminergic treatment, delusions and disease duration.

### Fronto-temporal disconnection linked to iPH

Clinical iPH vs. uPH differences were extended by our MRI findings. Resting-state functional connectivity within a defined PH network(29) showed that iPH-PD patients displayed lower fronto-temporal (IFG-pMTG) connectivity and variance differences in the right hemisphere. Similar changes in PH network connectivity were observed in psychotic patients with schizophrenia related to the loss of agency(55) or auditory-verbal hallucination(56). More generally, these findings align with the disconnection hypothesis, proposing lower connectivity for patients with behavioural and cognitive deficits, as shown in PD(57), Alzheimer’s disease(58) and schizophrenia(59). Moreover, although iPH-PD and uPH-PD did not differ in cognitive scores, overall IFG-pMTG connectivity was correlated with general performance in posterior cognitive tasks. Thus, the lower one’s IFG-pMTG functional connectivity, the stronger their posterior-cortical cognitive impairment. The posterior-cortical PD-CRS subscale is sensitive to cognitive decline (encompassing higher-order processes of semantic naming, visuospatial performance, affected in PD dementia) and VH (60), whereas the fronto-subcortical subscale measures attentional and executive processes (affected earlier in PD)(61). Linking iPH and posterior cognitive function to altered IFG-pMTG connectivity in the present sample is thus consistent with previous work associating structured VH to posterior cognitive impairments and compatible with our proposal that iPH may represent a non-visual ‘structured minor hallucination’ between uPH and other minor hallucinations and structured VH(9,60,62), although more longitudinal work is needed.

### Self-related and social mechanisms in iPH

More recently, PH has been interpreted as a hallucination of somato-motor origin, based on psychological intimacy with the presence(1), its location in peripersonal space(63), movement mirroring(28), as well as PH induction via conflicting self-related somato-motor signals in healthy subjects(30). The self-related origin of PH was further confirmed in PD patients, found to be more sensitive to the robotic induction, especially those with prior minor hallucinations(29). In the present patients we replicated past results of higher sensitivity following conflicting somato-motor stimulation(29,30,64) and found that iPH-PD versus uPH-PD did not differ in riPH sensitivity, showing that both groups share the self-related somato-motor origin of PH.

However, Fénelon and others suggested an alternative, social account of PH(4). Given that many of their PD patients reported iPH of specific identities of other people, authors suggested that PH results from interference with posterior brain areas of social perception and autobiographical memory networks in temporal cortex. We found such posterior regions for iPH in the present fMRI findings: namely pMTG (encompassing pSTS), a key region involved in social perception of biological motion(65), dynamic face expressions (particularly in the right-hemisphere)(66) and theory of mind (ToM)(67). It was previously also demonstrated as a modality-independent person-identity hub integrating identity information via Wernicke’s area and fusiform face area connections(68).

However, the present data show that almost all iPH were of first-degree proximity (Fig.1), revealing that only the closest others were experienced as iPH. Accordingly, we argue that a more specific social mechanism is necessary to account for iPH, involving regions implicated specifically in the processing of individuals of first-degree closeness(69,70) (e.g., pMTG and the default mode network are both modulated by subjective self-other proximity(71,72)). Significant others have also long been considered as one’s extended self(73) in memory(74) and perceptual studies(75,76). Integrating somato-motor and social accounts, we argue that iPH results from neurodegenerative interference with anterior somato-motor (self-related) as well as posterior cognitive (social) brain mechanisms, with uPH-PD lacking such posterior involvement. We also note that both systems are closely connected. Combined with recent evidence suggesting that PD patients exhibit deficits in minimal social cognition(77,78) and higher-order representation of other’s beliefs (ToM)(79) we speculate that iPH-PD vs. uPH-PD patients might exhibit stronger social deficits.

### Study Limitations

Although larger than previous work on PH, the present study is limited by its relatively small sample size. Extensive longitudinal studies should corroborate the present findings. Our sample’s insufficient posterior-cortical subscale variability could be the reason for our lack of group difference in cognitive performance. Furthermore, our resting-state FC analysis was limited to the PH network. Future work should address potential interactions with other regions or networks, previously associated with hallucinations. Finally, we only performed a somato-motor self-related paradigm (riPH task) but did not compare group’s social perception(64,80), which future work should quantify.

## Data Availability

All data produced in the present study are available upon reasonable request to the authors.

## Acknowledgment

The authors would like to thank all Parkinson’s disease patients for their participation, as well as clinicians Dr. Paul Krack, Dr. Julien Bally, Dr. Vanessa Fleury, Dr. Judit Horvath and Dr. Benoît Wicki for help with recruitment of PD patients.

## Author Roles

**Neza Vehar** – Research project: Conception; Organization and Execution. Statistical analysis: Design and Execution. Manuscript preparation: Writing of the first draft, Review and Editing.

**Jevita Potheegadoo** – Research project: Conception; Organization and Execution. Manuscript preparation: Reviewing and Critique. Resources, Supervision, Project administration.

**Nathalie Heidi Meyer** – Statistical analysis: Design, Execution. Manuscript preparation: Reviewing and Critique. Resources and Supervision.

**Léa Florence Duong Phan Thanh** – Research project: Execution (Data collection)

**Cyrille Stucker** – Research project: Execution (Data collection).

**Marie Elise Maradan-Gachet –** Research project: Execution (Data collection).

**Herberto Dhanis** – Research project: Execution (Data collection), Statistical analysis: Pre-processing of fMRI data.

**Lucas Burget** – Statistical analysis: Pre-processing of fMRI data.

**Sara Stampacchia** – Statistical analysis: Execution; Review and Critique. Manuscript preparation: Reviewing and Critique. Resources and Supervision.

**Fosco Bernasconi** – Research project: Conception. Statistical analysis: Design; Review and Critique. Manuscript preparation: Reviewing and Critique. Supervision.

**Olaf Blanke** – Research project: Conception; Organization. Statistical analysis: Review and Critique. Manuscript preparation: Writing of the first draft; Reviewing and Critique. Resources, Supervision, Project administration and Funding acquisition.

NV: 1A, 1B, 1C, 2A, 2B, 3A, 3B

JP: 1A, 1B, 1C, 3B

NHM: 2A, 2B, 3C

LFDPT: 1C

CS: 1C

MEM: 1C

HD: 1C, 2C

SS: 2B, 2C, 3C

FB: 1A, 2A, 2C, 3B

OB: 1A, 1B, 2C, 3A, 3B

## Ethical Compliance Statement

The study was approved by the Cantonal Ethics Committee of Geneva (Commission Cantonale d’Ethique de la Recherche sur l’Être Humain), protocol number 2019-02275. All patients consented to participation and signed a written consent form. We confirm that we have read the Journal’s position on issues involved in ethical publication and affirm that this work is consistent with those guidelines.

