## Supplemental information for "Parkinson’s disease progression and fronto-temporal disconnection relates to identity of minor hallucinations"

**Running title: Minor hallucinations and disease progression**

*Neza Vehar[1], Jevita Potheegadoo[1], Nathalie Heidi Meyer[1], Léa Florence Duong Phan Thanh[1], Cyrille Stucker[1], Marie Elise Maradan-Gachet2,4, Herberto Dhanis[1], Lucas Burget[1], Sara Stampacchia[1], Fosco Bernasconi[1], Olaf Blanke[1,3]*

[1]Laboratory of Cognitive Neuroscience, Neuro-X Institute, School of Life Sciences, Ecole Polytechnique Fédérale de Lausanne (EPFL), Geneva, Switzerland.

[2]Department of Neurology, Inselspital, University Hospital Bern, University of Bern, Bern, Switzerland.

[3]Department of Clinical Neurosciences, Geneva University Hospital, Geneva, Switzerland.

[4]Graduate School for Health Sciences, University of Bern, Bern, Switzerland.

**Keywords**: presence hallucination, self-other, phenomenology, resting-state fMRI, social familiarity

**Corresponding Author:**

Olaf Blanke

Bertarelli Chair in Cognitive Neuroprosthetics

Neuro-X Institute

School of Life Sciences

Campus Biotech

Swiss Federal Institute of Technology

Ecole Polytechnique Fédérale de Lausanne (EPFL)

CH – 1012 Geneva

### Supplemental Methods

The analysis was performed using R Studio's emmeans (1), lme4 (2), lmerTest (3) and ordinal (4) packages. The brain visualisation of PH network was produced using BrainNet Viewer (5).

#### *PH Questionnaire*

The Task 1 PHQ consisted of the 7 items, rated on the scale from 0 (Not at all) to 6 (Very much), presented in a randomized order between conditions. The items which were analysed below are listed in (6).

#### *Analysing resting-state functional connectivity of PH network*

Standard CONN preprocessing ("default_mnifield") pipeline was used (7). The results of the post-hoc analyses were corrected for multiple comparisons, using p-FDR<0.05 significance cutoff in R (fuzzySim library (8)).

**Table 1.** Regions-of-Interest coordinates for PH network taken from Bernasconi et al., 2021, (transposed bilaterally) in the Standard MNI space.

| **Brain region** | **MNI coordinates** | | |
|---|---|---|---|
| | **X** | **Y** | **Z** |
| Right IFG | 51 | 18 | 29 |
| Left vPMC | -53 | 1 | 37 |
| Right pMTG | 54 | -54 | 0 |

*Abbreviations: MNI – Montreal Neurological Institute, IFG – Inferior Frontal Gyrus, vPMC – ventral Premotor Cortex, pMTG – posterior Middle Temporal Gyrus.*

The results of the post-hoc analyses were corrected for multiple comparisons, using p-FDR<0.05 significance cutoff in R (using fuzzySim library (8)).

### Supplemental results:

#### *Robot-induced PH: Other PHQ items*

Ordinal regression mixed effects models were used to establish whether PHQ items differed between conditions and groups, with the following structure:

*PHQ item rating~Group*Delay+(1|patient_ID)*

For **self-touch** item, there was no main effect of group (OR=0.92, 95% CI[0.23,3.72], p=0.904) or delay condition (OR=1.09, 95 % CI[0.58,2.04], p=0.794), but we found a significant interaction group*condition (OR=0.47, 95% CI[0.24,0.91], p=0.026), implying the delay does not affect the groups equally, with higher ratings following asynchronous stimulation rather than synchronous stimulation for uPH-PD, but the reverse for iPH-PD.

For feeling like the patient was touched by someone else (**other-touch item**), there was no main effect of group (OR=0.90, 95% CI[0.31,2.55], p=0.837), delay condition (OR=1.84, 95% CI[0.96,3.51], p=0.067), or their interaction (OR=0.86, 95% CI[0.47,1.57, p=0.615).

Comparing **loss of agency item**, we found a significant main effect of delay (OR=1.96, 95% CI[1.01,3.77], p=0.045) with higher item scores in the asynchronous rather than synchronous condition, but no significant main effect of group (OR=1.01, 95% CI[0.40,2.52], p=0.990) or group*delay interaction (OR=0.98, 95% CI[0.55,1.77], p=0.960).

Investigating the PHQ item related to feeling of another minor hallucination termed the passage hallucination (**passage item**), the clmm model produced a warning ("Hessian is numerically singular, relative convergence criterion was unmet). To address model convergence failure, we opted for a simpler linear mixed effects model (lmer: passage ~ group * delay + (1 | patient_id)) and found statistically nonsignificant main effect of group (β=0.07, 95% CI[-0.21,0.35], p=0.605), delay (β=-0.03, 95% CI[-0.31,0.25], p=0.827) and their interaction (β=-0.20, 95% CI[-0.48,0.08], p=0.161).

Sensations item of the PHQ did not reveal a significant main effect for group (OR=0.70, 95% CI[0.35, 1.38], p=0.302), delay condition (OR=1.34, 95% CI[0.77,2.30], p=0.297) or their interaction (OR=1.16, 95% CI[0.69,1.97], p=0.570).

Two control questions were also posed in the PHQ: the front PH and the three bodies item, where no delay differences were expected, given that the stimulation does not normally result in such phenomenology and could be an indication of reporting bias. Lmer models had to similarly be adopted for the two control items due to convergence issues (estimated using REML and nloptwrap optimizer). We found that **the front PH item scores** did not significantly differ for group (β=0.06, 95% CI[-0.29,0.41], p=0.729), condition of delay (main effect: β=0.04, 95% CI[-0.14,0.21], p=0.676) or for the group*delay interaction (β=-0.03, 95% CI[-0.21,0.14], p=0.696).

Similarly, we found no significant main effect of group (β=-0.27, 95% CI[-0.67,0.14], p=0.193), condition of delay (main effect: β=0.02, 95% CI[-0.02,0.06], p=0.324) or for the group*delay interaction (β=-0.02, 95% CI[-0.06,0.02], p=0.324) for **the three bodies item** scores.

#### *Functional connectivity of PH network compared between groups*

We have conducted post hoc comparisons of 15 PH network edges between iPH-PD and uPH-PD patient groups with independent t-test, corrected for multiple comparisons (false detection rate (p-FDR)). All the edges but the edge between right-hemispheric IFG to pMTG, failed to survive the correction for multiple comparisons (Table 2) and were not investigated further for its relation to cognition.

**Table 2.** PH network edges' functional connectivity compared with the group contrast (iPH-PD vs uPH-PD).

| **Edge name** | **Estimate** | **SE** | **df** | **t-ratio** | **p-value** | **p-FDR adjusted** |
|---|---|---|---|---|---|---|
| R-IFG_R-pMTG | -0.163 | 0.05 | 236 | -3.254 | 0.001 | 0.020* |
| L-IFG_R_pMTG | -0.110 | 0.05 | 236 | -2.202 | 0.029 | 0.215 |
| L-vPMC_R_vPMC | -0.086 | 0.05 | 236 | -1.720 | 0.087 | 0.434 |
| L-IFG_L_pMTG | 0.077 | 0.05 | 236 | 1.534 | 0.126 | 0.474 |
| L-vPMC_L-pMTG | 0.053 | 0.05 | 236 | 1.067 | 0.287 | 0.767 |
| L-IFG_R-vPMC | -0.051 | 0.05 | 236 | -1.024 | 0.307 | 0.767 |
| R-IFG_L-vPMC | -0.033 | 0.05 | 236 | -0.668 | 0.505 | 0.884 |
| R-IFG_R-vPMC | -0.028 | 0.05 | 236 | -0.568 | 0.570 | 0.884 |
| L-vPMC_R-pMTG | -0.027 | 0.05 | 236 | -0.538 | 0.591 | 0.884 |

| | | | | | | |
|---|---|---|---|---|---|---|
| R-vPMC_L-pMTG | 0.025 | 0.05 | 236 | 0.492 | 0.623 | 0.884 |
| R-IFG_L-pMTG | -0.023 | 0.05 | 236 | -0.457 | 0.648 | 0.884 |
| L-IFG_L-vPMC | -0.014 | 0.05 | 236 | -0.286 | 0.775 | 0.969 |
| R_IFG_L-IFG | -0.010 | 0.05 | 236 | -0.192 | 0.848 | 0.969 |
| L-pMTG_R-pMTG | 0.006 | 0.05 | 236 | 0.120 | 0.904 | 0.969 |
| R-vPMC_R_pMTG | 0.001 | 0.05 | 236 | 0.025 | 0.980 | 0.980 |

*Abbreviations: R – right, L – left, IFG – Inferior Frontal Gyrus, vPMC – ventral Premotor Cortex, pMTG – posterior Middle Temporal Gyrus.*
